# Association of climatic determinants with Type 1 and Type 2 Diabetes worldwide: Night length and photoperiod variation linked to T1D and sunshine to T2D

**DOI:** 10.1101/2024.10.02.24314765

**Authors:** Julien Lacouchie-Payen

## Abstract

Nearly 500 million individuals are affected by diabetes worldwide. This very high prevalence is combined with a North-South gradient and a seasonality of diagnostics which all suggest the role of climate in diabetes etiology. However, only little is known about the impact of climate on diabetes. This article aims to understand the association of climatic variables with type 1 and type 2 diabetes (T1D and T2D) for 72 countries worldwide (1989-2021). T1D is, on average, more prevalent at extreme latitudes whereas T2D prevalence is higher near equator (*P* < 0,001). Sunshine, temperature, solar irradiance and daylength (photoperiod) are negatively associated with T1D prevalence and positively associated with T2D in simple regression (*P* < 0,001). Multicollinearity of climatic variables is considered as a challenge, and it is assessed with VIF and optimized with multiple regression. After adjustment, only photoperiod is associated with T1D prevalence (r^2^=0,45) and sunshine with T2D prevalence (r^2^=0,48). T1D monthly incidences are approximated with a cosine regression (RR=1,53) which is significantly associated with photoperiod along the year in Europe (*P* < 0,05). The relation between photoperiod and T1D has never been reported before in an ecological study and a short review is developed in the discussion.

---

The prevalence of diabetes is rising worldwide with more than half billion diabetic individuals affected today. This prevalence is expected to rise in the future decades ^1^ and this increase will have significant impact on individuals and health systems.

Diabetes is mainly represented by Type 1 Diabetes (T1D) and Type 2 Diabetes (T2D). T1D is more incident among young individuals and requires a life-long insulin treatment ^1,2^ whereas T2D reaches rather adults with several risks factors including overweight and physical inactivity ^1–3^.

For T1D, numerous studies have shown a spatio-temporal (latitude and season dependant) gradient of its incidence for different countries and regions in the world ^4–6^. Those seasons and geographic gradients are arguments for the role of climate in diabetes etiology. In some research, T1D distributions have been attributed to temperature ^7–11^ or ultraviolet B irradiance and vitamin D status ^5^ but it remains unclear which factor is the most associated. We also do the hypothesis that day length extremum would be associated with diabetes, indeed several studies have reported associations between circadian rhythm and diabetes ^12^ but no studies have shown this relation on ecological and world level.

In parallel, ecological research interested by climate and T2D are almost absent, even if it is, by far, the most frequent diabetes. Clinical evidence showed that temperature increased the severity of symptoms ^10,13^ and that month of patient birth or seasonal episodes of hyperglycemia could be associated with T2D risk ^14^. Those studies give argument for climate-dependent distribution of T2D, to our knowledge, only few studies are addressing this question. It should also be noted that studies are sometimes inconclusive ^15,16^.

More importantly, global warming will affect earth climate in the next decades. Climate change may modulate diabetes prevalence, as well as people wellbeing and public health measures ^17^ it is another reason why the relation climate-diabetes should be better understood today.

Therefore, this article will study both T1D and T2D diabetes and their associations with several climatic factors (temperature, sunshine, irradiance, daytime) at the global scale giving more clues about the environmental factor contributing to diabetes. Explaining models will be proposed regarding the results and literature.

## METHODS

### Diabetes data

Diabetes national prevalence for adults (20-79 years old, age-adjusted) and T1D national case number come from the International Diabetes Federation Atlas, 10^th^ edition^1^. The 68 countries/regions selected were chosen for the high quality of diabetes data^17^ and because all meteological data were also available for those countries. The total population included in this study accounts for approximately 5.420.183.000 individuals ^19^.

Between 85 and 95% of total diabetes prevalence is composed by T2D ^1,20^, therefore total prevalence are used as estimates of T2D prevalence, as already done in other studies ^20^. When available T1D prevalence data from diamond studies were used ^21^. For the analysis of T1D seasonality, incidence data (boys and girls, 1989-2008) came from a pan-national expert group (Eurodiab) focusing on T1D incidence found in 22 European countries and were graciously given by Dr. Patersson ^22^.

### Climatic data

Countries latitudes are centroid latitudes^23^. The sun-related parameters come either from equations (daytime, total irradiance) or ground stations (temperature, sunshine). Datas for Health expenses per capita are expressed in current US dollars ^19^, this variable is used as control for confusion factors regarding healthcare level.

The daytime (D) equation comes from the National Oceanic and Atmospheric Administration (NOAA) based on the method of Jean Meuus ^24^ and gives the length of the day from sunrise to sunset (in minutes) such as:

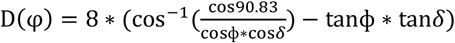

*φ* and *δ* are respectively the latitude of the corresponding country (°) and the sun declination (°). Reversely, average yearly night length (Fig.2) is obtained by subtracting D to the calendar length of the day (24 hours). It is worth mentioning that high average for yearly daytime for a country also means high variations in the daytime during the year. Repartition between night and day length is called photoperiod in the rest of the article.

Irradiance (I) is calculated at noon on the top atmosphere on the date of the winter solstice for each country; I(*φ*) was obtained with the formula: *I*(ϕ) = 1367 * cos(ϕ ± 23.5). It comes from total irradiance equation ^25^ and is used by other research about irradiance and health ^5^.

The ground measured temperatures (monthly max and min) were retrieved from the Global Historical Climatology Network Daily (GHCND) from the NOAA database for the years between 1975 and 2010 ^26^. Ground measured sunshine durations were gathered from the United Nation database for the years between 1961 and 2015. Using more than 30 years of data allows us to study climate as defined by World Meteorological organization ^27^. Meteorological stations were chosen based on their proximity to countries’ centroids.

Monthly irradiance (all sky surface downward irradiance) used in seasonality study are obtained from the NASA Langley Research Center POWER (project from NASA Earth Science Directorate Applied Science Program). Data are accessible online.

### Min, Max, Average temperature and sunshine

Association between diabetes and average, maximum or minimum monthly temperature (or average, max or minimum sunshine) have been evaluated. Average temperature and minimum monthly sunshine were the most associated parameters. Our understanding is that sunshine measurements are more sensitive to the minimum value (near the burning threshold of the pyranometer).

### Meteorological definitions

Solar irradiance indicates total solar radiation reaching one latitude on top atmosphere. Here, it is calculated for the winter solstice, and it is an indicator of solar intensity at this moment as it has been done in previous reference article^28^. Instead of using complex correction or extrapolation to deduce ground irradiance, we used sunshine which measures the duration of a solar beam with a ground instrument (pyranometer with a burning threshold near 120 W/m2). It indicates direct solar radiation (both in intensity and volume) is based on empiric ground measurement and accounts for the cloudiness. Daytime indicates the duration of the day from sunrise to sunset, it both measures direct and diffusive sunlight and puts emphasis on duration of sunlight rather than only intensity.

### Statistics and data processing

The regressions are based on polynomial regressions (2^nd^ order) for Fig. 1; on linear regression for Fig. 2, Tab. 1 and 2; and on cosine regression for Fig. 3. Multiple linear regression (MR) is used in Tab. 1 and 2, the optimized model includes only 2 variables and is based on a sequential MR giving the most efficient model. Variance inflation factor (VIF) is calculated to control the risk for multicollinearity (Tab. 3). All data are processed with R (3.6.2) and figures are done in Libre Office.

**Figure 1.**
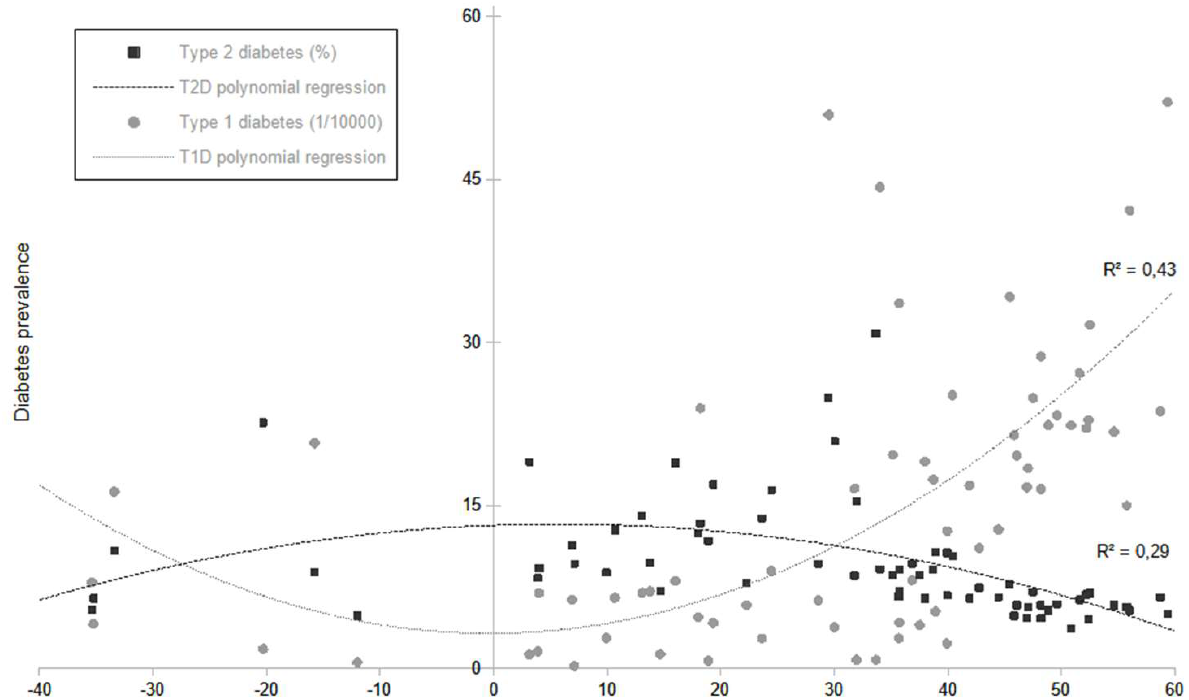
Geographic latitudes are associated with T1D (1/10000; ?) or T2D prevalence (%; ■) for 68 countries worldwide (2021). Polynomial regression (2^nd^ order) fits both distribution, r^2^=0,43 for T1D, r^2^=0,29 for T2D (p<0.001). List of the countries ranked by latitude : New Zealand, Argentina, Australia, Chile, Mauritius, Brazil, Peru, Malaysia, Colombia, Maldives, Sri Lanka, Venezuela, Costa Rica, Trinidad et Tobago, Barbados, Thailand, Phillipines, Sudan, Virgin Islands (U.S); Puerto Rico (U.S), Dominican republic, Mexico, Honk Kong, Oman, United Arab Emirates, India, Kowait, Egypt, Israel, Jordan, Pakistan, Morocco, Cyprus, Japan, Algeria, Iran, Rep. of Korea, Greece, Portugal, United States of America, China, Turkey, Spain, Italy, Bulgaria, Romania, Canada, Croatia, Slovenia, Switzerland, Moldova, Hungary, Slovakia, Austria, France, Luxembourg, Belgium, United Kingdom, Poland, Netherlands, Germany, Lithuania, Russian Fed., Denmark, Estonia, Sweden, Finland

**Figure 2.**
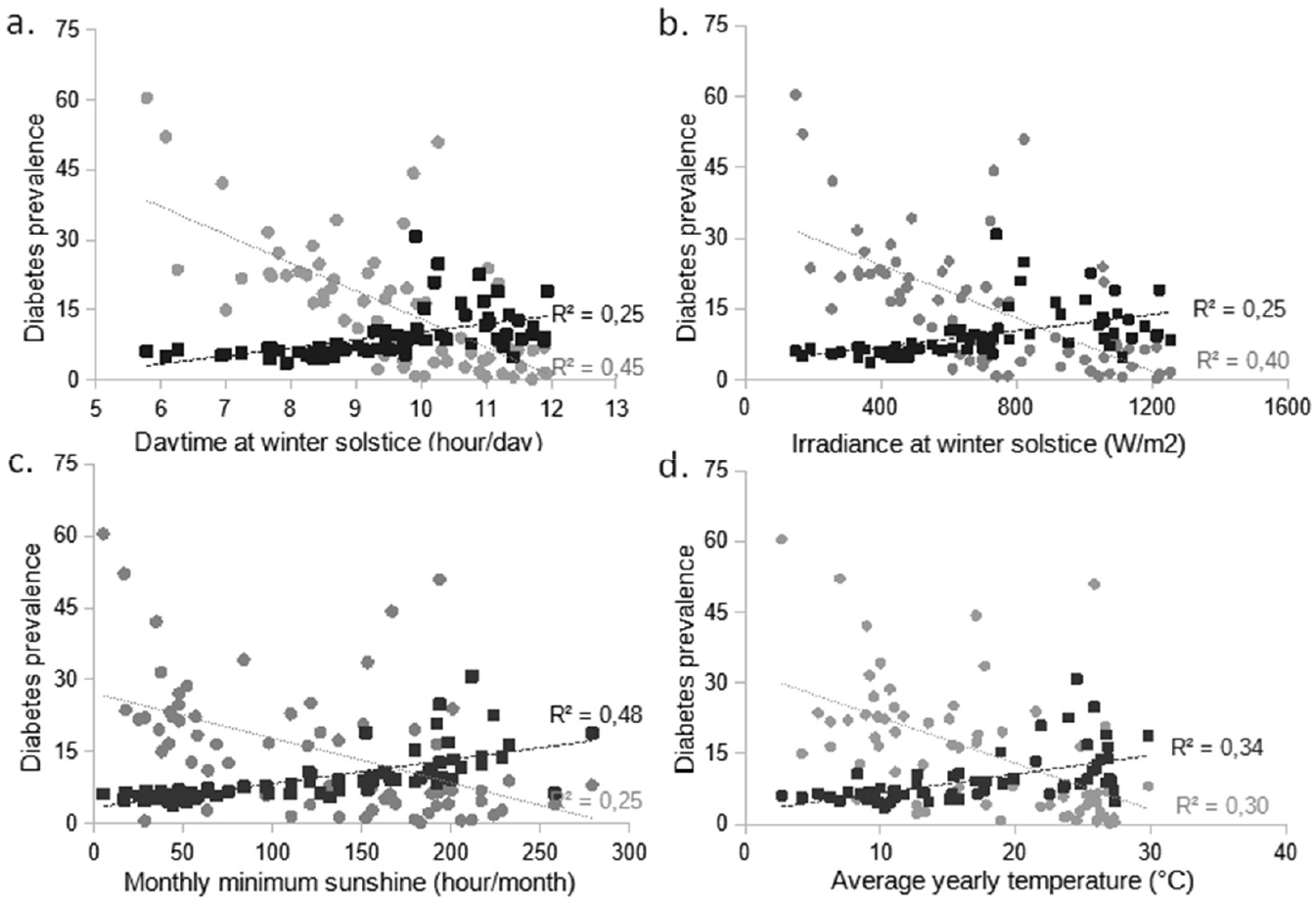
Average daytime at winter solstice (a), irradiance at winter solstice (b), monthly minimum sunshine (c), yearly average temperature (d) are associated with T1D (1/10000; ?) or T2D (%; ■) for 68 countries worldwide. Associations are based on simple linear regressions and diabetes prevalence are significantly associated for each variable (p<10.e-4).

**Figure 3.**
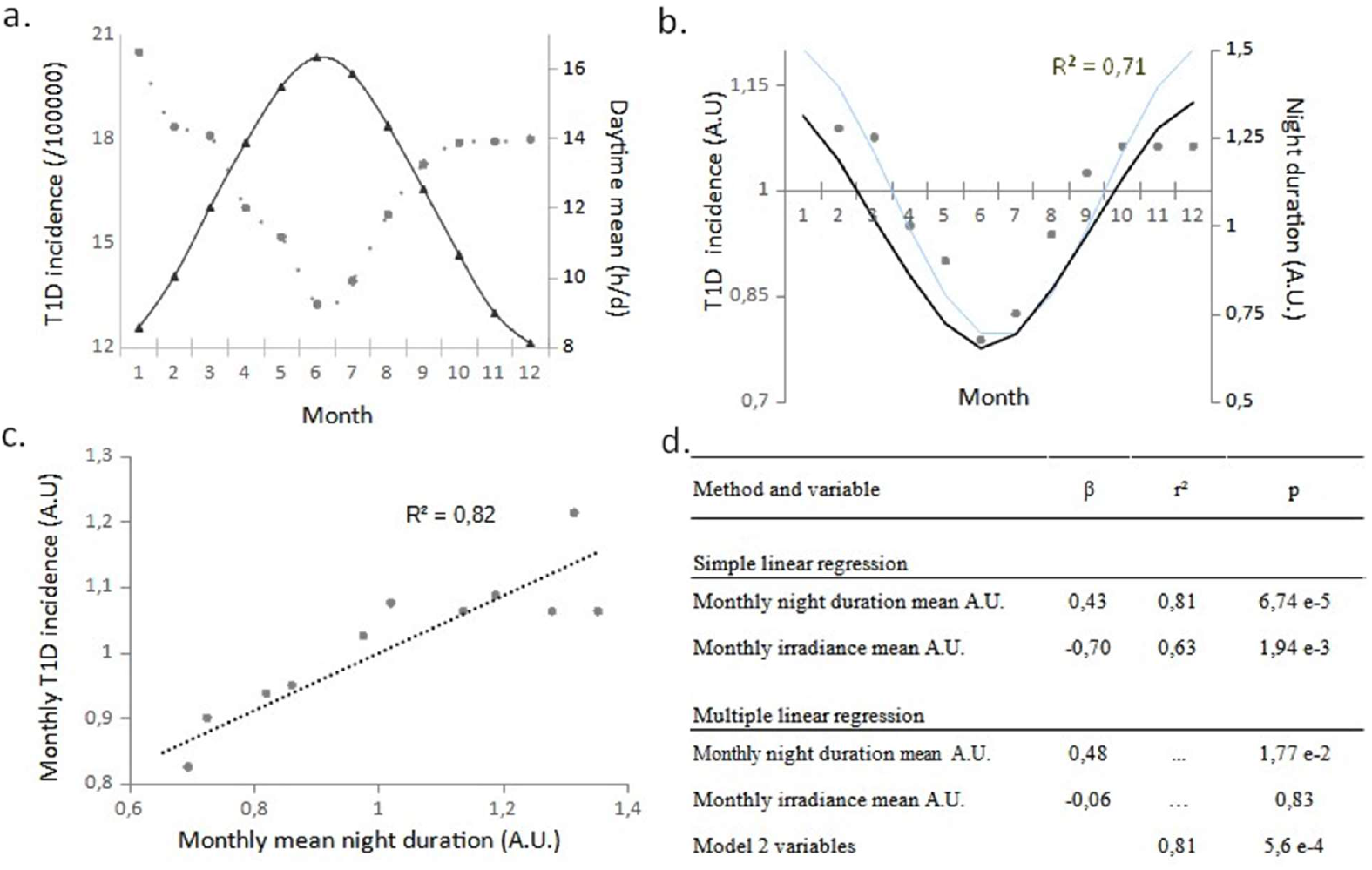
T1D monthly incidence is significantly associated with photoperiod along the year in Europe a. Monthly T1D incidence for 23 Eurodiab centers (1/100 000; ?) and photoperiod (continuous line) variation along the year b. normalized T1D incidence approximated with cosine regression based on Broohart method (grey line, r^2^=0.71, p<0.001; RR = 1,53) and normalized night length (black line) along the year c. monthly T1D incidence plotted against monthly mean night duration with simple linear regression (r^2^=0.82, p<0.001) d. Simple and multiple regression for Irradiance total or/and night duration explaining T1D incidence. Incidence diabetes data are pooled results for 84 658 T1D cases from 19 European countries diagnosed between 1989 and 2013. Normalization (A.U) is done by centering the data on the average of the dataset. Daytime, night duration and irradiance are based on 50°N latitude, the average latitude of the 23 centers.

### Seasonality study

The seasonality study employs the improvement of the geometric method of Edward brought by Brookhart and Rothman ^29,30^.

In summary, the seasonality of the monthly count of T1D cases are tested by comparing the distribution of diabetes data against the variance of a homogeneous (non-seasonal) distribution (χ^2^, 2 df). Because this test was significant, a non-homogeneous Poison process is applied and monthly T1D cases are estimated by the function *f* and can be expressed as follow:

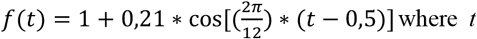

is the numeric value of the month (Jan=1, Dec=12). To confirm the goodness of the fit a χ^2^ test (11 df) is done.

## RESULTS

### Latitudes are associated with T1D or T2D prevalence with opposite trends

T1D prevalence tends to be higher at extreme latitude whereas T2D prevalence is more important near tropics (p<0.001, 67 dof). From our knowledge, it is the first time that a relation between latitude and T2D prevalence is presented. Relations between prevalence and latitude gives a rational for studying associations between sun-related parameters and diabetes.

### Sun-related parameters are associated with diabetes prevalence

T2D prevalence (Fig. 2 and Tab. 1) is most associated with monthly minimum sunshine (r^2^=0.48, *P*=7,45e-11) followed by average yearly temperature (r^2^=0.34, *P*=2,19e-7), both of which are positive associations. An increase of 1 hour of monthly minimum sunshine is associated with an increase of 0,05 % [0,04;0,06] of T2D prevalence. It should also be noted that an increase of 1 °C is associated with 0,4 % of T2D prevalence [0,26;0,54], this result could be considered for global warming and its potential impact on human health.

**Table 1.** T2D prevalence (%) - Simple and multiple linear regression with sun-related variables.

| Method and variable | $\beta$ | CI | $r^2$ | p |
| --- | --- | --- | --- | --- |
| Simple regression |  |  |  |  |
| Monthly minimum sunshine (hour/month) | 0,05 | [0,04 ; 0,06] | 0,48 | 7,45 e-11 |
| Average yearly temperature (°C) | 0,40 | [0,26 ; 0,54] | 0,34 | 2,19 e-7 |
| Irradiance at Winter solstice (W/m<sup>2</sup>) | 0,008 | [0,005 ; 0,012] | 0,25 | 1,36 e-5 |
| Daytime at Winter solstice (hour/day) | 1,71 | [0,98 ; 2,44] | 0,25 | 1,77 e-5 |
| Health expenses per capita (\$) | -0,0009 | [-0,0015 ; -0,0003] | 0,12 | 3,88 e-3 |
| Multiple regression - total <sup>a</sup> |  |  |  |  |
| Monthly minimum sunshine (hour/month) | 0,05 | [0,03 ; 0,07] | ... | 6,34 e-5 |
| Model 5 variables |  |  | 0,50 | 1,89 e-8 |
| Multiple regression - optimized <sup>b</sup> |  |  |  |  |
| Monthly minimum sunshine (hour/month) | 0,05 | [0,04 ; 0,06] | ... | 3,76 e-9 |
| Health expenses per capita (\$) | -0,0003 | [-0,0008 ; -0,0002] | ... | 0,27 |
| Model 2 variables |  |  | 0,49 | 4,00 e-10 |

<sup>a</sup> Multiple regression - total includes all independent variables (5), only sunshine is significantly associated in this model.
<sup>b</sup> Multiple regression - optimized includes only two variables maximizing the determination coefficient.
Confidence interval (CI), 95 %.

For T1D prevalence (Tab. 2), daytime at winter solstice is the most associated variable (r^2^=0.45, *P*=3,19e-10) followed by irradiance (r^2^=0.39), those are negative associations. Curves shows that daytime and diabetes are particularly associated for higher latitude (over 30° or less than 10 hours daytime at winter solstice).

### Photoperiod is significantly associated with T1D prevalence and sunshine is significantly associated with T2D prevalence once adjusted for other sun-related parameters

MR models (Table 1 and 2) explain up to 50 % of diabetes prevalence by environmental factors. Once adjusted with sunshine for T2D or day length for T1D no other variables are significantly (*P*<0,05) associated with diabetes prevalence. Nevertheless, each variable is independently associated with diabetes prevalence in simple linear regression models. This contradictory result may be explained by a certain level of multicollinearity relation that is studied hereafter (Tab. 3). Those results suggest that models for T1D must include photoperiod whereas models for T2D should include sunshine other parameters may add precision on case by case.

**Table 2.** T1D prevalence (1/10000) - Simple and multiple regression with sun-related variables.

| Method and variable | $\beta$ | CI | $r^2$ | p |
| --- | --- | --- | --- | --- |
| Simple regression |  |  |  |  |
| Daytime at Winter solstice (hour/day) | -6,03 | [-7,63 ; -4,43] | 0,45 | 3,19 e-10 |
| Irradiance at Winter solstice (W/m<sup>2</sup>) | -0,03 | [-0,04 ; -0,02] | 0,39 | 7,47 e-9 |
| Average yearly temperature (°C) | -0,98 | [-0,62 ; -1,34] | 0,3 | 1,31 e-6 |
| Monthly minimum sunshine (hour/month) | -0,09 | [-0,13 ; -0,05] | 0,25 | 1,54 e-5 |
| Health expenses per capita (\$) | 0,003 | [0,001 ; 0,004] | 0,16 | 7,92 e-4 |
| Multiple regression - total <sup>a</sup> |  |  |  |  |
| Daytime at Winter solstice (hour/day) | -12,33 | [-20,72 ; -3,94] | ... | 5,53 e-3 |
| Model 5 variables |  |  | 0,5 | 2,05 e-8 |
| Multiple regression - optimized <sup>b</sup> |  |  |  |  |
| Daytime at Winter solstice (hour/day) | -9,09 | [-12,89 ; -5,29] | ... | 1,43 e-5 |
| Average yearly temperature (°C) | 0,67 | [-0,09 ; 1,43] | ... | 8,68 e-2 |
| Model 2 variables |  |  | 0,48 | 6,89 e-10 |

<sup>a</sup> Multiple regression - total includes all independent variables (5), only daytime at winter solstice is significantly associated in this model.
<sup>b</sup> Multiple regression - optimized includes only two variables maximizing the determination coefficient.
Confidence interval (CI), 95 %.

**Table 3.** VIF between independant variables evaluating collinearity. Formula for VIF = 1/(1-r^2). Data are the same as those used in Table 1, Table 2 and Figure 2.

| Variance inflation factor (VIF) | Average yearly temperature (°C) | Minimum monthly sunshine (h/y) | Daytime at WS (h/d) |
| --- | --- | --- | --- |
| Irradiance at WS (W/m2) | 7,38 | 2,24 | 29,43 |
| Average yearly temperature (°C) | - | 2,84 | 5,82 |
| Minimum monthly sunshine (h/y) | - | - | 2,37 |

### Variation of T1D monthly incidence is significantly associated with photoperiod along the year in Europe

The association between lower daytime at winter solstice and higher T1D prevalence for 68 countries (tab. 2) translates a spatial distribution of the disease because daylength is itself a function of latitude and solar declination. This relatively high correlation (r^2^=0,45) could suggest causality. This possibility would be reinforced if there is temporal relation temporality between daytime and T1D incidence variation along the year, this relation is assessed in Fig. 3.

The pooled data for European T1D incidence (both gender) are not homogeneously distributed along seasons (*P*<0.001). Cosine regression is applied for the distribution of monthly T1D incidence in Europe (r^2^=0.71, *P*<0.001) and correlates well with night length variation along the year (Fig. 3) and gives a seasonal relative risk of 1,53 for T1D incidence along the year. Monthly T1D incidence and night length are minimal in June (Summer solstice) and are maximal between December and January (near winter solstice) (Fig. 3 a).

In Fig. 3 b, a linear correlation between monthly night duration and monthly T1D incidence confirms a strong temporal correlation between both T1D and photoperiod along the year in Europe (r^2^=0.81, *P*<0.001), the power of the association should be confirmed by increasing the statistical power.

### Multicollinearity is a challenge for ecological studies regarding climatic solar variables

Variance inflation factor (Tab. 3) aims to study collinearity between independent variables to confirm Tab. 1 and 2 results. Collinearity is viewed as significant if 10 > VIF > 5 ^31^. Temperature is collinear with Irradiance and daylength (resp. VIF = 7,38 and 5,82). Sunshine is not or only weakly collinear with the other variables. Daytime and irradiance are strongly collinear (VIF = 29,43) however the Daytime includes irradiance (power) as well as the dimension of time (day length or photoperiod) which could explain why irradiance is not associated with T1D prevalence once adjusted for daylength.

## DISCUSSION

This study shows, for the first time, an association between T2D prevalence and latitude (Fig. 1). T2D tends to be maximum near equator and to be lower at higher latitude. For T1D, some studies have reported lower incidence near the equator and higher incidence at the pole ^4,5^. Those results are reinforced here with prevalence data (Fig. 1). Unlike incidence studies, prevalence distributions suggest link to longer term trends such as evolutive, climatic or solar trends ^25^. Several studies reported a seasonality for T1D occurrence throughout the year and this is also shown in fig. 3 for European countries ^32–34^. Taken together, these results indicate that sun-related parameters modulate diabetes occurrence even if there is no consensus over the most important environmental parameter ^4,5,8,9,32^. This article aims to understand better the relation between climate and diabetes.

Dahlquist and Mustonen ^32^ showed that diabetes incidence for Swedish children was independently associated with low sunshine hours and low mean temperature. In the present article, monthly minimal sunshine hours were not associated with T1D prevalence once adjusted for mean temperature (Tab. 2). This result is in accordance with the study of Waenbaum ^9^ which found an association between T1D and temperature but not with sunshine. This is questioning how much sunshine dependent vitamin D pathway is important in type 1 diabetes etiology at high latitude.

From our knowledge, it is also the first time that an association between T1D prevalence and Day length at Winter solstice is reported (r^2^=0.45, Tab. 2). Indeed, sunshine or temperature association becomes insignificant after adjustment for daytime (*P* < 0.01). These results are in accordance with Patterson research that sunshine hours and average temperature have only little short-term influence on T1D incidence ^8^. It is also in agreement with the association between UV and T1D ^5^ as Daytime period is both an indicator of sunlight duration and intensity. In our results, photoperiod explains 45% of T1D prevalence and its spatial distribution (Fig. 1, Fig. 2, Tab. 2). Accordingly, several high incidences of T1D are found in the north of China ^34^.

To test the relation between T1D and daytime, the relation between monthly incidence and monthly average night duration was also investigated for 19 European countries (Fig. 3). T1D incidence is estimated with a circannual rhythm approximated with cosinus regression (r^2^=0,71) and night duration strongly correlates with T1D monthly incidence (r^2^=0,82) suggesting a potent role for photoperiod variation along the year in T1D etiology. Night duration remains the only variable significantly associated with T1D incidence once adjusted for irradiance (Fig 3 d). This result confirms the association between T1D incidence and Daytime/Night duration on the temporal scale for European countries. It should be noted that T1D incidences of Children in Shangai and Japan seem to follow a similar pattern along the year ^6,35^.

The positive association between sunshine and T2D (Tab. 1) is unexpected because sunshine will normally increase vitamin D synthesis and high level of vitamin D normally decreases T2D risk ^36^. Other factors might explain this result, very high sunshine may decrease physical activity during those periods as reported in Qatar ^37^. Moreover, Fonseca and al. ^38^ have already reported frequent vitamin D deficiency in Saudi Arabian women even if the country is very sunny, possibly because of custom and sedentary lifestyle. Interestingly, national distribution of T2D in India have higher prevalence in South India ^39^ which is in agreement with a positive association between T2D and sunshine.

High temperature reduces brown adipose tissue (BAT) catabolism, glucose combustion ^10^ and insulin sensitivity ^40^ increasing the risk for hyperglycemia and diabetes. Higher temperature may also decrease the outdoor physical activity over a certain threshold as it was observed for a senior cohort in Japan ^41^. Moreover, once adjusted with sunshine (Tab. 1), temperature is not associated with T2D prevalence. An explanation could be that sunshine generally precedes temperature increase. This result makes sunshine an interesting positive predictor for T2D prevalence (r^2^=0,48, *P*=7,45 e-11).

To sum up, the present model shows that sunshine is relevant for predicting T2D prevalence whereas photoperiod variation is important for studying T1D prevalence and incidence. Possible explanations and mechanisms for this new association will be reviewed hereafter.

T1D etiology is usually explained by the expression of genetic susceptibility triggered by an environmental factor ^42,43^. Environmental factors that have already been studied for T1D etiology are viral or bacterial infection ^44,45^, vitamin D status ^5,36, 42,46^-both of which have not being proved as direct causal factors ^42^ and more recently sleep disorder or circadian and circannual rhythm have also been studied ^47–49^.

Circadian rhythms are near 24 hours metabolic cycles related to daily activity and those are often connected to circannual rhythm which depends on yearly season, night-day length and biologic constraints ^50,51^. Those rhythms are based on ‘Zeitgeber’ stimuli (such as light/dark) or time giver stimuli that are giving environmental clues on the time of the day. Day length and photoperiodism gives the most noise free signal to predict time of the year to anticipate and adapt the metabolism to the milieu ^50^.

Light-Dark zeitgeber is picked up by the retina and passes through the Supra Chiasmatic Nucleus (SCN) ^12^. SCN is the main circadian clock of the human body and sends message to the pineal gland and the secondary circadian clocks. Those are specialized cells with autonomous but adaptable cycles and include the insulin producing Beta cell ^52,53^. MLN is the main hormone produced by the pineal gland during dark phase ^50,54^ and is responsible for the synchronization of circadian rhythms ^55–57^. Seasons modulate MLN secretion in plasma, maximal mean is reached during the winter solstice in December and the minimum during summer solstice in Jun ^58–61^ and higher MLN concentrations are associated with higher latitude during winter ^51,62-64^.

Numerous studies have reported that MLN reduces insulin levels ^65,66^ or glucose tolerance in animals and human ^67–70^. Indeed, knock-out of circadian genes (which are dependent on MLN) in pancreatic β cell changes melatonin-insulin equilibrium and leads to diabetes ^53,71^. Moreover, Melatonin Receptor 1 B (MTNR1B) variants account for 30 % of genetic susceptibility of T2D for European descendants ^72^ and involve a loss of function of the receptor ^73^ and an increase in fasting glucose ^74^. Those patients produce less MLN and insulin at the basal level or after glucose challenge (OGTT) ^75,76^ suggesting a causal link. Very recently, Daher et al. ^77^ also found mutations in an MLN receptor (MTNR1A) associated with renal dysfunction in T1D patients advocating for the role of MLN signaling in T1D complications.

The variation of daytime across latitude and along season (at high latitude) modulates the Light-Dark cycle of individuals (Fig. 3). Photoperiod variation observed at high latitude changes the stimulation of SCN and various studies have confirmed the variation of MLN linked to latitude and season ^62,63^. This variation might cause a circadian disruption as it is observed in several diseases ^12,48,49,54^. The Glacer study has also shown an association between several circadian genes’ variants (CYR1 and 2 or MTNR1B) and higher glycemia in a season dependent manner for non-diabetic people in Sweden ^78^ suggesting an early role of photoperiod response on diabetes onset.

It remains that T1D is highly dependent of genetic and most frequent genes associated with T1D are related to HLA and DR3, DR4-DQ8 variants ^79–81^ Those are coding for Major Histocompatibility Complex 2 (MHC 2) and are responsible for microbial antigen presentation but are also involved in the presentation of self-peptides in autoimmunity especially in T1D ^81,82^. Several autoantibodies have been found in T1D patients specially insulin autoantibody^82^. Current T1D etiology explains that self-antibodies are triggering an autoimmune destruction of beta cell in Langerhans islet from pancreas resulting in deficiency of insulin, hyperglycemia and onset of T1D ^42, 81-83.^ In humans, IgG1 is the most precocious and frequent antibody found among insulin autoantibody in human patient ^84–86^. Interestingly, IgG are upregulated by short day/long night duration in several animals ^87–89^.

Moreover, MLN acts as modulator of the immune system ^90–95^. Piolli and his colleagues showed that MLN increased MHC 2 expression on splenic macrophage and antigen presentation which in turn increases proliferation of T-lymphocyte ^94^.

Interestingly, T1D associated genes are often also associated with sleep disorders or circadian rhythm disturbances. For example, MHC and HLA-DQ loci are implicated in narcolepsy ^96–97^ as well as in diabetes ^79^. Several other genes have been found to be associated with diabetes such as LMO7 or PTPN22, IL-2/IL-2R or FoxP3 and those genes have been linked to sleep disorders or circadian disorders ^80,88,89,99,100^.

For example, LMO7 variants are also risk factors for long sleep duration ^101^ and is regulated by day length in other species ^102,103^. PTPN22 is also frequently reported in T1D genes studies, it is a phosphatase involved in the blockage of lymphocytes activation which is important for autoimmune regulation ^104,105^ and PTPN22 is also expressed deferentially in transcriptomic analysis regarding circadian changes ^106^.

Numerous studies have reported sleep disorders in T2D and there is a growing body of evidence for the involvement of sleep disorders in T1D ^107–110^. Most common sleep disturbances found in T1D are sleep restriction ^110^ and sleep quality problems ^107–109^. Moreover, social jet lag, a kind of circadian misalignment (changes in sleeping schedule because of social activity) is also associated with a significative reduced glycemic control for T1D patients ^111–112^.

In summary, the association between T1D and night length, sleep disorder and circadian misalignment as well as the important decrease in MLN concentration in T1D patient seems that studying the effect of photoperiod, circadian rhythm and sleep-awake cycle on immune regulation of self-antibodies before the onset of T1D would be interesting.

## Limit of the study

This article aims to give a world portrait of diabetes-climate distribution. However, some regions are less represented data were absent or of low quality. In parallel, certain countries with good datasets but relatively small populations are highly represented such as several islands or european countries. Also, some countries with high population (China, India etc.) have the “same weight” than smaller country in this study.

Studying climate and several solar variables is often associated with multicollinearity problems. Most articles already published about diabetes (or other disease) epidemiology use only one or two climatic variables which is a risky approach as it is highly prone to multicollinearity, it is also a rationale for the present research. Nevertheless, because of the multicollinear nature linked to solar variables the present results should be taken with caution and confirmed with experimental setting and metanalysis.

Last and not least, circadian rhythm studies have sometimes conflicting results regarding the effect of MLN even in GWAS study^113^. The use of a nocturnal animal (*Mus musculus*) may not be ideal to infer function in diurnal humans. Those problems give more value to epidemiological and ecological studies regarding photoperiod and circannual rhythm for human.

## Strength of the study

Near 5,5 billion people are included in this study, which is an important part of world population.

The climatic variables that have been chosen are simple and robust. Little or no transformation of the meteorologic data has been used. This should increase understanding as well as the reliability of the study. Ground station and astronomically calculated variables are easily accessible.

The use of several variables allows to control multicollinearity problems which is often not considered by other ecological study regarding human health. Several diabetes datasets have been used (IDF, Diamond) for T1D or T2D prevalence and T1D incidence. Their association with climatic variables is consistent and it increases the power of the present research. Also, T1D and T2D are studied simultaneously, which is not the case of all diabetic study. Several clinical, epidemiological and experimental articles are in accordance with our results.

## Conclusion

This research gives an overview of the association between several climatic factors and diabetes. It uncovers the important association between photoperiod and T1D as well as the association of sunshine with T2D. The positive association between temperature and diabetes could also be considered for prediction of T2D regarding climate change and global warming. Critical moments of photoperiod should be determined with clinical setting to understand better the relation photoperiod with T1D but also to act on specific moments in individuals’ lives in order to prevent diabetes. At the era of climate change and global warming, this study gives data to assess diabetes-climate association and ideally predict diabetes prevalence.

## Data Availability

The article is currently under review by a journal and not available for now

## Acknowledgment

I would like to share my sincere thanks to Dr Sharif Mohr for his time and experienced counsels, to Dr Chris Patterson for sharing its diabetes incidence data and his advice and to Dr Mathieu Ferron for the initial and interesting work on diabetes in his lab. The author has no relation of interest to declare. This work began with a Molson-Bombardier grant and a merit grant from the faculty of medicine (university of Montreal).

